# Optimal time to return to normality: parallel use of COVID-19 vaccines and circuit breakers

**DOI:** 10.1101/2021.02.01.21250877

**Authors:** Michael B. Bonsall, Chris Huntingford, Thomas Rawson

## Abstract

By January 2020, the COVID-19 illness has caused over two million deaths. Countries have restricted disease spread through non-pharmaceutical interventions (e.g., social distancing). More severe “lockdowns” have also been required. Although lockdowns keep people safer from the virus, they substantially disrupt economies and individual well-being. Fortunately, vaccines are becoming available. Yet, vaccination programs may take several months to implement, requiring further time for individuals to develop immunity following inoculation. To prevent health services being overwhelmed it may be necessary to implement further lockdowns in conjunction with vaccination. Here, we investigate optimal approaches for vaccination under varying lockdown lengths and/or severities to prevent COVID-19-related deaths exceeding critical thresholds. We find increases in vaccination rate cause a disproportionately larger decrease in lockdowns: with vaccination, severe lockdowns can reduce infections by up to 89%. Notably, we include demographics, modelling three groups: vulnerable, front-line workers, and non-vulnerable. We investigate the sequence of vaccination. One counter-intuitive finding is that even though the vulnerable group is high risk, demographically, this is a small group (per person, vaccination occurs more slowly) so vaccinating this group first achieves limited gains in overall disease control. Better disease control occurs by vaccinating the non-vulnerable group with longer and/or more severe lockdowns.

## Introduction

The emergence of the SARS-CoV-2 virus in late 2019 has had, subsequently, devastating consequences across the globe, with all countries reporting levels of virus infection affecting public health responses. Since the emergence of this novel coronavirus, the policies implemented (in part using insights from previous pandemics) have focused on non-pharmaceutical interventions (NPIs) and approaches such as travel bans, limited household mixing, stay-at-home orders (‘lockdowns’) and quarantines, social distancing and the closure and restriction of large mass gatherings. These NPIs require effective implementation, behavioural shifts and continued political and public support [1, 2] and have mitigated disease spread, levels of infection, rates of morbidity, and mortality [3, 4, 5]. While these measures have suppressed related COVID-19 deaths, NPIs have macroscale impacts on the economics of many nations [6, 7, 8], as well as, arguably, microscale implications, affecting the mental well-being of many individuals [9].

Although these non-pharmaceutical approaches have some success in reducing virus spread, they are not a sustainable strategy, and other treatment-based approaches are required [10]. Several candidate vaccines are in Phase III trials reporting efficacies from 70% to more than 90% [11]. At the end of 2020, two vaccines (Pfizer-BioNTech and Oxford-AstraZeneca) have been approved by regulators in some countries as safe for general use.

Even under the assumption that vaccines work as intended, the challenges now are three-fold. The first is to instigate mass vaccination programs. The second is to prevent health services being overwhelmed in the meantime, recognising that to vaccinate a nation takes time and that there is a lag of some weeks after inoculation before high immunity levels are achieved. The third issue is that many citizens are weary, and persuading them to follow restrictive rules while a vaccine appears imminent may be difficult. That is, many in society could be overly confident that the threat posed by the virus is reduced in light of the announcements of a successful vaccine, failing to recognise there remains a time lag before full levels of herd immunity are realized.

There is therefore a requirement to determine the optimal balance between five factors. These factors are: (1) total number of deaths (that in some situations be regarded as a proxy for healthcare capacity), (2) vaccination rate, (3) the level of lockdown restrictions impacting virus transmission rates, (4) the order in which different population groups are inoculated and (5) time spent in lockdown.

Here we solve aspects of this optimisation problem, based on predictions from a standard SIR (Susceptible, Infected and Recovered) model, modulated by a vaccination program. Specifically, we calculate for prescribed total number of deaths, achievable vaccination rates of different groups and transmission rates, the optimal policy of vaccination delivery amongst population groups so as to minimise time spent in lockdown. We introduce heterogeneity in terms of group structure, with a focus on three main population groups of vulnerable, front-line worker, and non-vulnerable. Our overarching aim is to demonstrate how vaccination and lockdowns can, and should, be used together to achieve optimal disease control. In the the next section we introduce the mathematical frameworks and methods of analysis. In the results, using numerical approaches and as parameterised for the UK epidemic, we illustrate how different levels of vaccination, lockdown severity and prescribed maximum level of disease determine a minimal lockdown length for a derived optimal strategy of vaccination across our three groups. In the discussion, we present our results in light of previous work on vaccination and non-pharmaceutical interventions.

## Mathematical Models

### Basic Unstructured Model

Mathematical models such as the SIR (Susceptible, Infected, Recovered) models are commonly used frameworks to describe disease transmission [12]. To explore the combined effects of vaccination and lockdowns on controlling infections, we begin by using a simple extension to an SIR framework in which vaccinated individuals (*V*) are accounted for separately from recovered (*R*) individuals. The dynamics for susceptible (*S*) individuals is such that the rate of change of the number of susceptibles decreases due to individuals becoming infected, is increased by any loss of immunity of people who have previously recovered, lowered by background death rate and lowered as people are vaccinated. Hence, this gives for the time evolution of *S*:

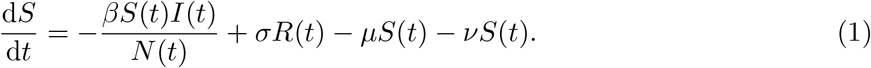

The dynamics for infected individuals are such that the number increases as susceptibles pass through an incubation period of length *τ* become infectious. The number of infected decreases as people either die from COVID-19, die naturally, or recover. Hence *I* follows:

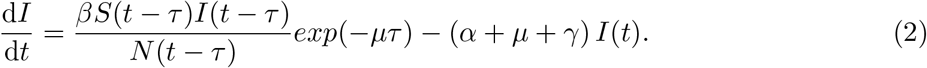

The dynamics for recovered individuals are such that the number increases as people recover from infection, or decreases due to vaccination, non-COVID-19-related death, or loss of immunity, giving:

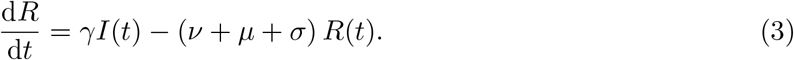

Finally the dynamics for the total number of vaccinated individuals is dependent on the vaccination rate, lowered only by a background death rate, and therefore there is an assumption of no immunity loss for those inoculated and no onward transmission of virus once individuals are vaccinated. Hence:

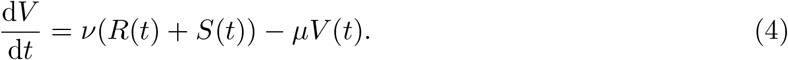

In equations (1)-(4), parameter *β* is the disease transmission rate, *σ* the loss of immunity, *µ* is the background death rate, *ν* is the vaccination rate (acting on both susceptible and recovered individuals), *τ* is the incubation window (and *exp*(−*mutau*) is the survival rate through the incubation window), *α* is the disease induced death rate, and *γ* is the disease recovery rate. We introduce circuit breakers as a reduction in transmission such that the value of *β* reduces to a range of values. The total number of people is *N* (*t*) = *S*(*t*) + *I*(*t*) + *R*(*t*) + *V* (*t*).

The optimal control problem (Appendix 1) is formulated as finding an optimal vaccination rate (*ν*(*t*)) under different levels of lockdown restriction (i.e. different levels of *β*) within a time interval [0,*T*] so as to minimize ‘costs’ of vaccination and disease-induced deaths above a threshold *Z*:

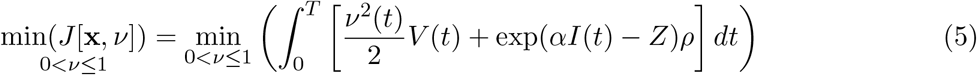

with control inequality constraint 0 *≤ ν ≤* 1, subject to the system of governing differential equations (**x** = [*S, I, R, V*]^⊤^) and initial conditions **x**(*t*(0)) = **x**_**0**_, where *T* is the length of epidemic (and not the lockdown period) and *ρ* is a scaling constant. The increasing quadratic ‘costs’ associated with vaccination assumes that vaccination becomes increasingly difficult as the number of daily vaccinated individuals increases - this is one way to describe ‘vaccination demand’ (see below for other approaches to this on the disease dynamics). The function exp(*αI*(*t*) − *Z*) describes the difference between the number of disease-induced deaths at time point *t* and a considered threshold value *Z*. Raising this difference to an exponent ensures that the contribution of (*αI* − *Z*) is small if *αI* < *Z* and that (*αI* − *Z*) contributes greatly to the cost functional *J* (equation (5)) when *αI* > *Z*. This means that deaths, above a threshold *Z*, are increasingly penalised, corresponding to exceeding a normal acceptable level of healthcare capacity. Here, *ρ* is a scaling constant that, within the objective functional, weights the relative contribution of the threshold mortality effects compared to vaccination ‘costs’ on the minimisation.

### Structured Model

We extend the model to incorporate demographic structure, corresponding to different groups of people. Individuals are classed as ‘front-line workers’ (*FR*), ‘vulnerable’ (*V*) individuals or ‘non-vulnerable’ (*NV*) individuals. Transmission could occur differently within and between groups, and we define *β*_*j,i*_ as the probability that an infected individual in group *i* infects a susceptible individual in group *j*. The force of infection (the *per capita* rate of infection) for the *i*^th^ group is then:

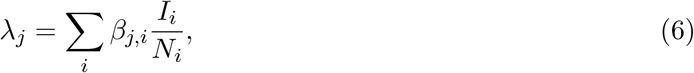

where 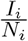 is the frequency-dependent transmission function in the *i*^th^ group. The expected probability that a susceptible individual (in group *j*) acquires an infection, from any source, is then the sum of the products of transmission rate and proportion of infected individuals in the *i*^th^ group, for all *i*.

The use of equation (6) allows us to retain the mathematical structure of the epidemiological dynamics of (1)-(4), now applied to the *i*^th^ group as:

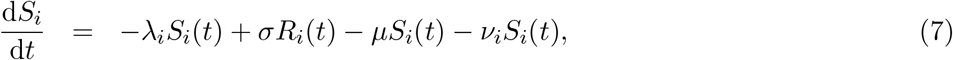

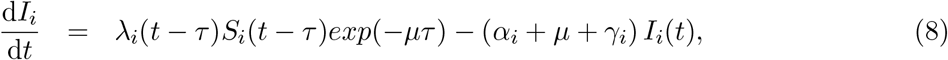

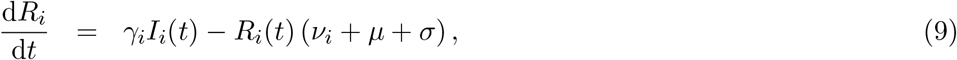

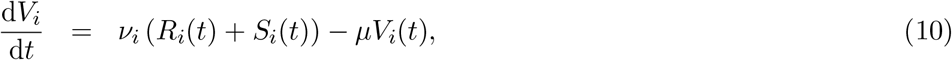

where vaccination rate (*ν*_*i*_), disease-induced death rate (*α*_*i*_) and recovery rate (*γ*_*i*_) are group-specific parameters. The rate of loss of immunity (*σ*), background death rate (*µ*), and virus incubation time (*τ*) are population-level parameters are instead independent of group structure. For each group *N*_*i*_(*t*) = *S*_*i*_(*t*) + *I*_*i*_(*t*) + *R*(*t*) + *V*_*i*_(*t*).

### Optimal vaccination

To investigate the optimal vaccination level under different levels of lockdown severity to keep disease-induced mortality below a critical threshold, we solve the optimal control problem defined by equation (5) following Pontryagin’s maximum principle [13]. This solution is for the unstructured model (equations (1)-(4), considering the population as a single cohort defined by *S, I, R* and *V* subgroups). Using the objective functional (equation (5)), adjoint equations, and control inequality constraint [14], optimal dynamic vaccination rates (*ν*(*t*)) are calculated for varying levels of reduced disease transmission (*β*). We use a modified Runge-Kutta method (a forward-backward-sweep algorithm [13]) to find the optimal outcome. The full details of the corresponding Hamiltonian and adjoint equations used to identify the optimum vaccination rate are presented in the Appendix.

### Vaccination, different cohorts and lockdowns

We also investigate the interplay between vaccination strategies amongst population groups and length of lockdowns, so as to achieve the optimal suppression of the virus (in terms of disease-induced mortality) for a variety of considered lockdown lengths and effectiveness, while keeping cumulative mortalities below a critical threshold. To do so we use the structured model (equations (7)-(10)). This model framework accounts for the three groups of front-line workers (*FR*), vulnerable (*V*) and non-vulnerable (*NV*), and was solved numerically over 150 days. The model was parameterized with parameters estimated to be appropriate to the UK (Tables 1 & 2). We calculate the optimum vaccination strategies across different population groups.

**Table 1:** Population-level parameters fixed across all groups.

| Parameter | Definition | Value | Source(s) |
| --- | --- | --- | --- |
| $\beta$ | transmission rate | 0.016 | [23] |
| $\mu$ | background mortality rate | $2.273 \times 10^{-5}$ | [27] |
| $\tau$ | incubation time | 5.1 | [28, 29] |
| $\nu$ | vaccine efficacy | 0.001 – 0.05 | this work |
| $\sigma$ | loss of immunity | (set to 0) | this work |

**Table 2:** Population-level parameters specific to the different individual groups.

| Front-line worker group |  |  |  |
| --- | --- | --- | --- |
| Parameter | Definition | Value | Source(s) |
| $\alpha$ | disease-induced death rate | 0.0032 | [30] |
| $\gamma$ | recovery rate | $\sim 0.1$ | [31, 32] |
| Vulnerable individual group |  |  |  |
| Parameter | Definition | Value | Source(s) |
| $\alpha$ | disease-induced death rate | 0.064 | [30] |
| $\gamma$ | recovery rate | $\sim 0.06$ | [32, 33] |
| Non-Vulnerable individual group |  |  |  |
| Parameter | Definition | Value | Source(s) |
| $\alpha$ | disease-induced death rate | 0.0032 | [30] |
| $\gamma$ | recovery rate | $\sim 0.1$ | [31, 32] |

### Optimal lockdown times

Importantly, we investigate the hypothesis that lockdowns can be used to support, in parallel, on-going vaccination programs in order to prevent infection rates crossing key thresholds. In particular, we consider how different strategies concerning the order in which groups vulnerable, front-line workers, and non-vulnerable are inoculated, for the same maximum infection rate thresholds, impacts on length of lockdown. Hence the optimum we search for is the shortest lockdown strategies for each threshold. Again, we solve the structured epidemiology model (equations (7)-(10)) numerically for different sequences of vaccination over 150 days. The chosen sequence is to deliver vaccine to first group for 30 days, first and second group from 31-60 days and all groups after 60 days. Critically, the vaccination rates, expressed as fraction of cohort vaccinated per day, are common to each group. This implies that as the vulnerable group is smaller, the number of people vaccinated per day is smaller, likely reflecting the actual situation. The optimal outcome in terms of group order vaccination strategy to achieve the shortest lockdown times, while maintaining cumulative mortalities below a critical threshold, is also determined for varying lockdown severities (i.e. levels of transmission reduction).

### Vaccination demand: declining vaccine uptake rates

A further component that we consider is vaccination elasticity. This process is used to describe how supply and demand varies given changes in (usually) price of a commodity [15]. In epidemiology, this idea of elasticity in demand has been linked to vaccine uptake and disease prevalence [16]. With inelastic demand in vaccine, as disease prevalence falls and assuming that vaccination levels can be maintained, then the public health benefits of vaccination (and the resulting herd immunity) can be maintained as infection levels fall. However if vaccine uptake is elastic with respect to disease prevalence, such that as disease prevalence falls individuals are less likely to take a vaccine, then the public health control of infections can be more challenging. Elasticity can prevent achieving the vaccination levels needed to reach the critical threshold for herd immunity.

Here we investigate the role of vaccine elasticity demand, via a time evolving function *ν*(*t*), on disease outcomes (in terms of cumulative levels of mortality) under different lockdown lengths and severities. Our working hypothesis, that we test, is that lockdowns can be used to offset vaccine elasticities to maintain and/or achieve virus control. We use numerical approaches to solve the full structured model over time (150 days) to find the optimal vaccination level, lockdown duration and severities that keep cumulative mortality below a critical threshold.

The code used for all numerical analyses and simulations is available at https://osf.io/xvunt/.

## Results

### Optimal vaccination

Solving the constrained optimisation problem (Appendix) shows that the optimal vaccination strategy is a function of the ratio between the susceptibles, recovered and vaccinated individuals:

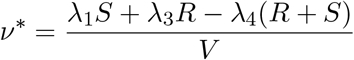

where *λ*_*i*_ are adjoint (Lagrange) multiplier variables associated with the state variables (*S, R* and *V*). While the state variables constrain the minimisation of the objective functional (5), these multipliers can be thought of as representing costs of violating the state variable constraints (Appendix).

Solutions for the optimal vaccination rate are shown in Figure 1. As planned, coupled with the threshold mortality condition, an optimal vaccination strategy can suppress the COVID-19 epidemic (Figure 1A). However, for increasing lockdown severity (20-80% reduction in transmission i.e. *β*), this can lead to the infection fading out without the characteristic epidemic growth curve associated with general S-I-R type dynamics.

**Figure 1:**
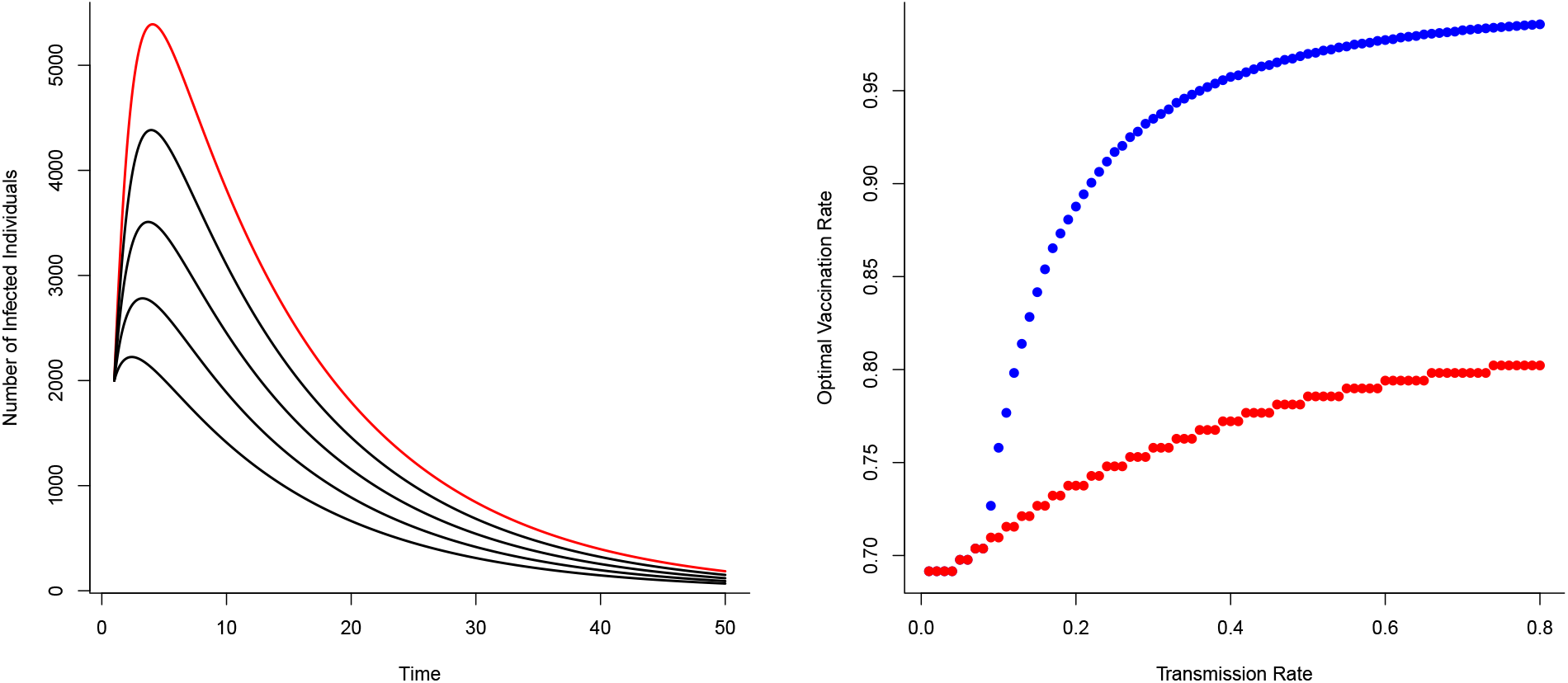
Optimal vaccination rate *ν*^*\**^ on disease control (from the unstructured S-I-R-V model) under a fixed critical threshold (*Z*) that instantaneous disease-induced deaths should not exceed. (A) The effects of reducing transmission (*β*) on disease dynamics for, in red, no reduction in transmission (*β* = 0.8) with progressive reductions (20%, 40%, 60% and 80%) in transmission shown in black (*Z* = 100). (B) Optimal vaccination rate for different transmission rates for different fixed critical threshold (*Z*) (blue points *Z* = 10; red points *Z* = 100). Optimal vaccination rate with low transmission (severe lockdown) reduces the potential for epidemic disease epidemics and minimize number of deaths. (Other parameters: *β* = 0.8. *α* = 0.01, *γ* = 0.1, *µ* = 0.001, *ρ* = 1.0).

With a canonical parameter set (Figure 1), optimal vaccination strategies that keep daily disease-induced mortality below a critical threshold (*Z* = 100) and can reduce peak numbers of infections by ∼ 74% (with just vaccination) through to ∼ 89% (with vaccination coupled with a 80% reduction in transmission rate), all compared to the no-control scenario.

The (optimal) vaccination strategy is influenced by the stringency of the mortality threshold (*Z*) (Figure 1B). As might be expected, a more stringent threshold necessitates higher levels of vaccination to achieve the expected level of control to ensure disease-induced mortality remains below that critical threshold. As also expected, decreasing potential disease transmission (e.g. through the use of NPIs) offsets the need for high levels of vaccination to achieve the necessary levels of disease control (Figure 1B). Notable is the strong non-linearity in Figure 1; where, as a result of reductions in transmission (i.e, effects of different lockdown scenarios), for different critical thresholds the optimal vaccination rate declines in different ways for different critical thresholds.

### Vaccination, different cohorts and lockdowns

Under the structured epidemiological dynamics (equations 7-10), cumulative levels of disease-induced mortality is influenced by the length of time in, and severity of, lockdown along with the vaccination rate (Figure 2). Increasing the vaccination rate decreases the need for longer lockdowns and, under high vaccine coverage, the necessity for these lockdowns. While still essential, lockdowns can instead be of short duration when partnered with a vaccination program. This is again to limit cumulative mortalities to different levels. Under lockdowns where the level of transmission is only reduced by 20% or 40%, cumulative mortalities due to the virus are expected to be excessively high unless vaccine coverage is also high. Under more severe lockdowns where transmission is reduced by 60% or 80% (Figure 2), shorter circuit breakers can be sufficient to limit disease-induced mortalities.

**Figure 2:**
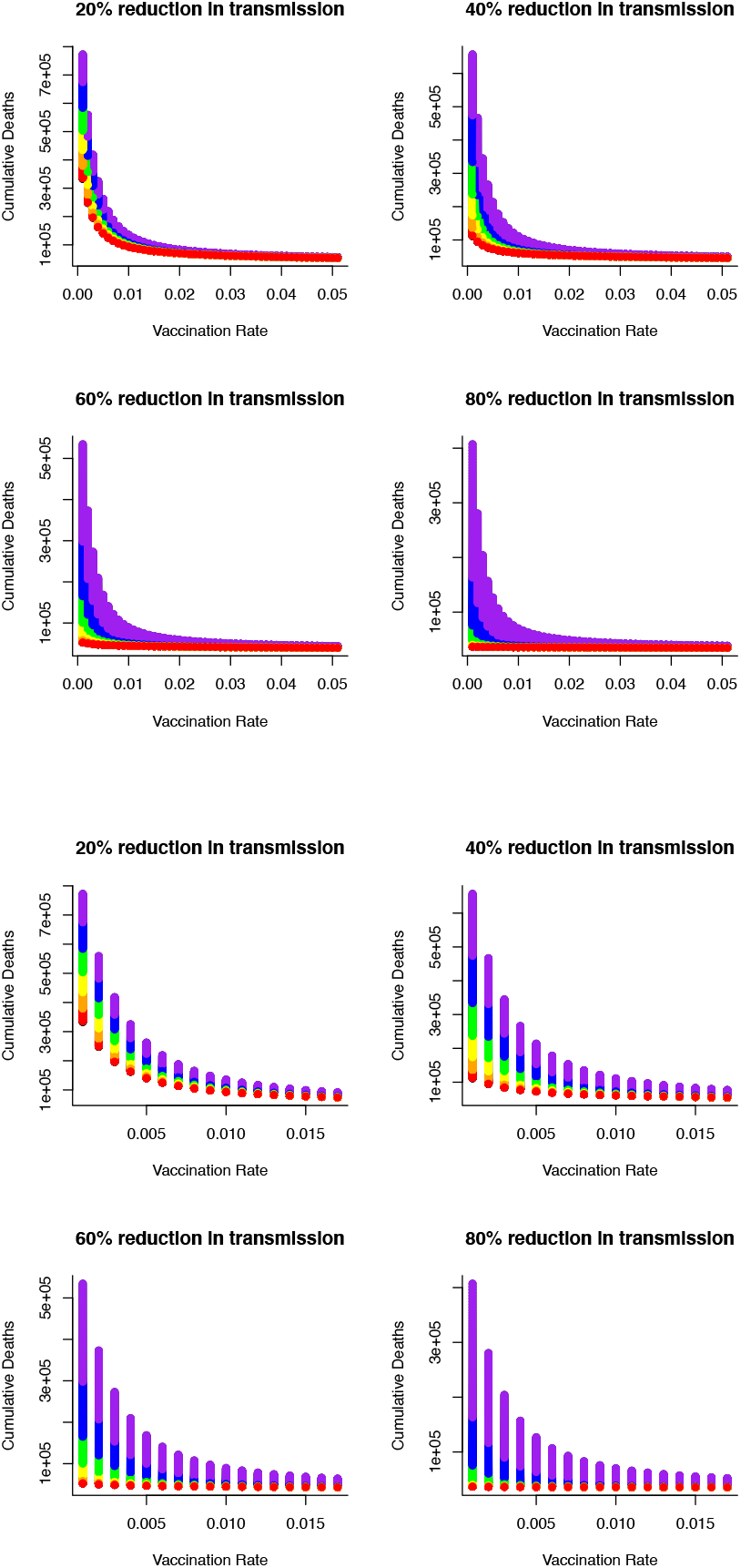
Combined effects of vaccination and lockdown durations (rainbow colours blue/purple short (15 days) through to red - long (95 days)) for different lockdown severities (where transmission is reduced by (a) 20%, (b) 40%, (c) 60% and (d) 80%) on cumulative deaths. For vaccination to be at all effective in reducing cumulative mortality (say 50K over 150 days) in the range of coverage anticipated (Figure B 0.006 - 0.0152) then lockdown severity needs to reduce transmission by at least 60% or lockdown durations need to be unduly long.

More specifically, for vaccination to be effective in reducing cumulative mortality (set here at ∼ 50, 000 over 150 days), and in the range of anticipated vaccination rate (0.006 - 0.0152), then lockdown severity needs to reduce transmission by at least 60% or lockdowns need to be unacceptably long and extend for more that 90 days. Furthermore, these sort of lockdowns or circuit breakers can be used to offset weakly efficacious vaccine rates to ensure that disease-related mortalities are kept in check below the (nominal) threshold of 50, 000 (Figure 3).

**Figure 3:**
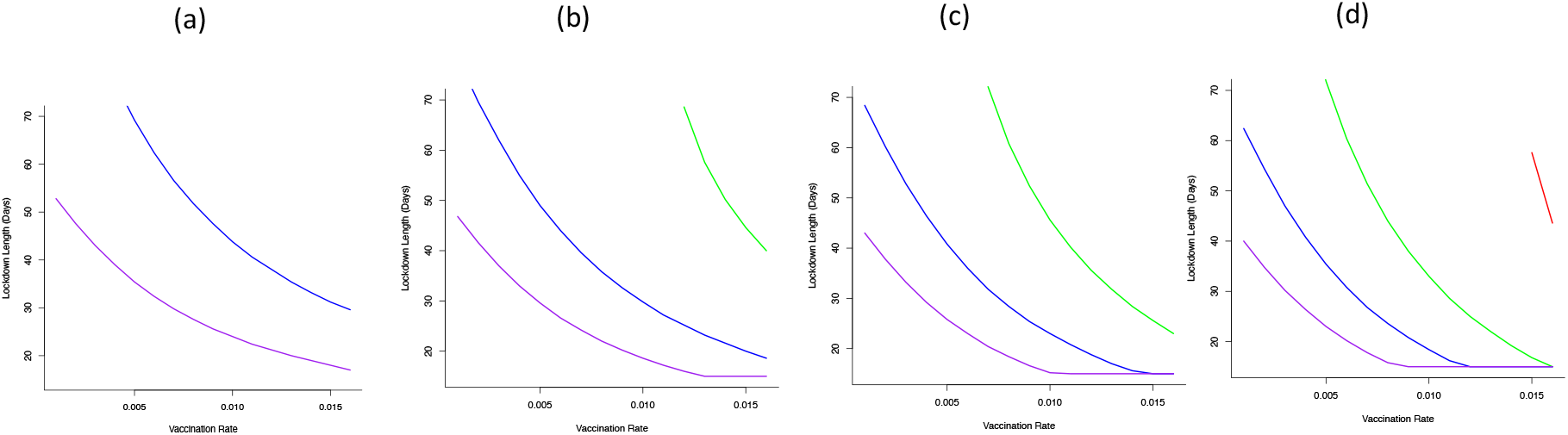
Optimal combination of vaccination rates (covering an expected vaccination rate range of 0.006 - 0.0152) and length of lockdowns to (under different severity to keep cumulative deaths below (a) 50K (b) 60K (c) 70K or (d) 80K for different lockdown severity (purple - 80% reduction in *β*, blue - 60% reduction in *β*, green - 40% reduction in *β*, red - 20% reduction in *β*). From (a) – 50K mortality threshold, for a weakly efficacious vaccination rate (0.005), depending on the severity of lockdown, lockdown durations could be 30 days (for 80% reduction in beta) or 70 days (for 60% reduction in beta). At this level of vaccination (0.005) and for a lockdown where *β* is only reduced by 20% or 40% it is not feasible to keep cumulative mortality below the 50K threshold.

To understand more fully vaccine delivery strategies between different parts of society, we begin by determining how focusing initial vaccine delivery on single groups affects the likelihood of keeping levels of mortality below the (optimal) threshold. If vaccines are delivered singly to vulnerable or key worker groups then lockdowns would still be necessary and they would need to be reasonably severe (> 60% reduction in transmission) to reduce cumulative mortality below key thresholds (Figure 4). Notable is that less severe lockdowns when vaccinating either of these two groups are not sufficient to keep cumulative mortality below a critical threshold. This key finding is critical, and requires consideration in light of the decision by many countries to vaccinate the most vulnerable first. The reason for this finding is due to the demographic differences between these two groups (where population sizes in these groups are relatively small) compared to the non-vulnerable group (which contains the majority of the population). For example, consider a vulnerable (V) group of 500,000 people, and a non-vulnerable (NV) group of 10,000,000 people. If we vaccinate roughly 5% of each group each day this would lead to vaccinating either 25,000 vulnerable people, or 500,000 non-vulnerable people, daily. Hence, even if the vulnerable people are an order of magnitude more likely to die of COVID-19, more lives will be saved by vaccinating the 500,000 non-vulnerable group. For some nations this scenario might be likely, if non-vulnerable people can be vaccinated (in units of people per day) at a speed that is an order of magnitude larger - for instance through mass vaccination centres that the vulnerable are unable to reach. In those circumstances, a more robust approach to achieve keeping mortality below the critical threshold would be to vaccinate across the non-vulnerable group and gain from the related shorter and/or less severe lockdown (Figure 4).

**Figure 4:**
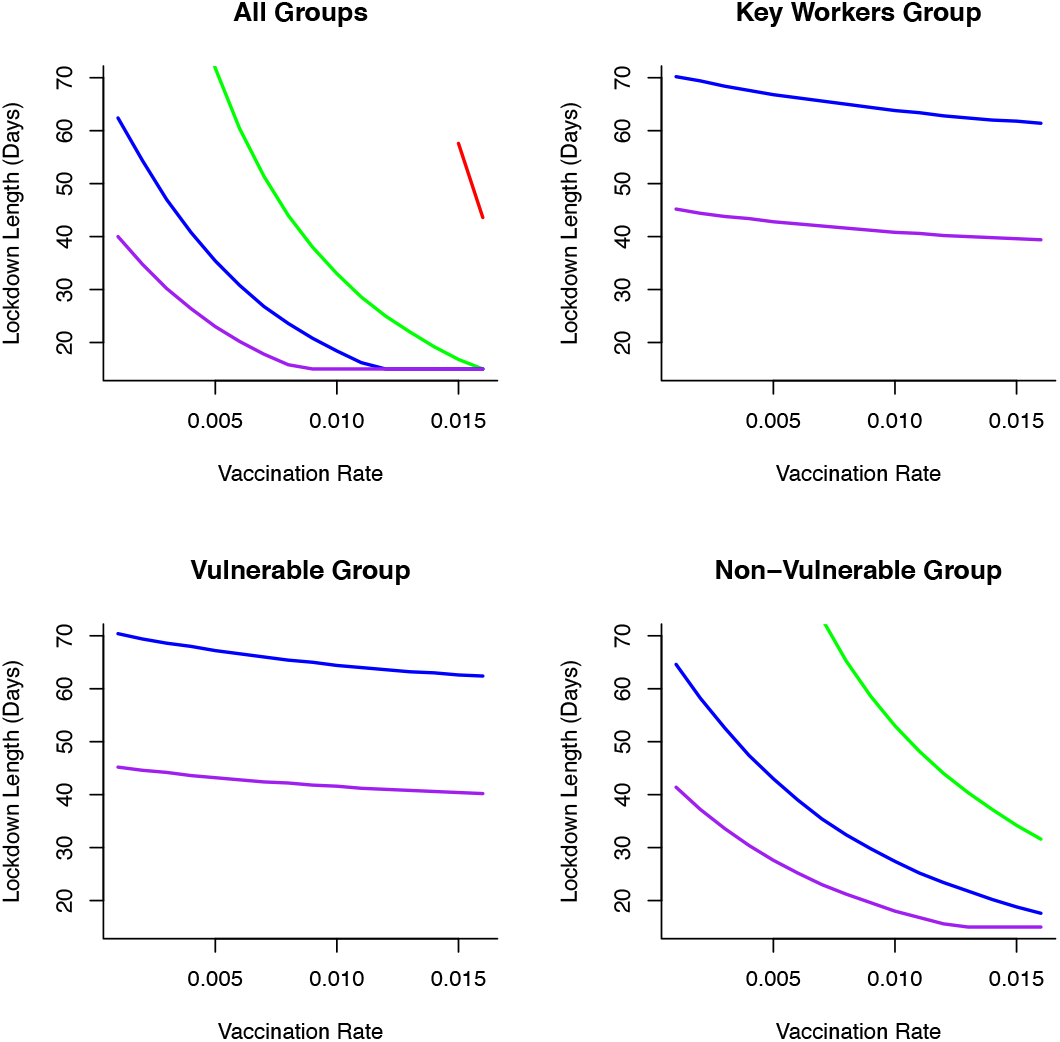
Combinations of vaccination across different groups: (a) all groups vaccinated (b) only key worker group vaccinated (c) only vulnerable group vaccinated (d) only non-vulnerable group vaccinated, lockdown duration (in days) and lockdown severities (in terms of reducing transmission-(purple - 80% reduction in *β*, blue - 60% reduction in *β*, green - 40% reduction in *β*, red - 20% reduction in *β*)) on keeping cumulative mortality less than 80K. Vaccination across the non-vulnerable group provides greater opportunities to keep mortality below critical threshold with lockdowns of short duration and/or less restrictive.

### Optimal lockdown times

Optimal outcomes, in terms of shortest possible lockdown times based on order of vaccination between cohorts, are shown in Figure 4 for different transmission reductions. This summary figure shows how different combinations of vaccination and lockdown interventions can lead to successful disease control (in terms of minimizing cumulative mortalities) (Figure 4). With low vaccination rate (e.g. *ν* = 0.005), and depending on the severity of lockdown, lockdown durations could be as short as 20 days (for a 80% reduction in *β*). Alternatively, a lockdown may be 30 days (for 60% reduction in *β*) or 50 days (for 40% reduction in *β*) for successful virus control. However, under this level of vaccination (*ν* = 0.005), it is simply not feasible for a lockdown where *β* is only reduced by 20% to keep cumulative mortality below the (nominal) 80, 000 threshold.

We illustrate how vaccinating different groups in different sequences can influence strategies to achieve the optimal outcome, and most notably impact the time required in lockdown. If a strategy is adopted to vaccinate different groups over sequential 30 day periods (and continuing this vaccination strategy once initiated for each group), then the choice of longitudinal sequences influences the choice of lockdown duration and/or severity to achieve optimal disease control. For the majority of sequences, moderate to severe lockdowns are needed to achieve the optimal goal of keeping cumulative mortality below a critical threshold (Figure 5). However, adopting a strategy in which the non-vulnerable group is vaccinated first, followed by the front line worker group followed by the vulnerable group (“NV-FR-V”; Figure 5) allows lockdowns of weaker severity (where disease transmission is reduced by only 20%), albeit these might be of longer duration, to be used to achieve the optimal goal. Apart from this sequence of vaccine delivery, there is again no feasible vaccination strategy where weak lockdowns (transmission is reduced by 20%) would allow mortality levels to be kept below 50,000 people. Targeting the non-vulnerable group in the first or second wave of vaccination allows a broad set of lockdown severity (40%-80% reduction in transmission) strategies to be implemented. If a strategy is adopted to target vulnerable and front line workers then it will require moderate to severe lockdowns (60%-80% reductions in transmission) to achieve the optimal outcome for disease control (Figure 5).

**Figure 5:**
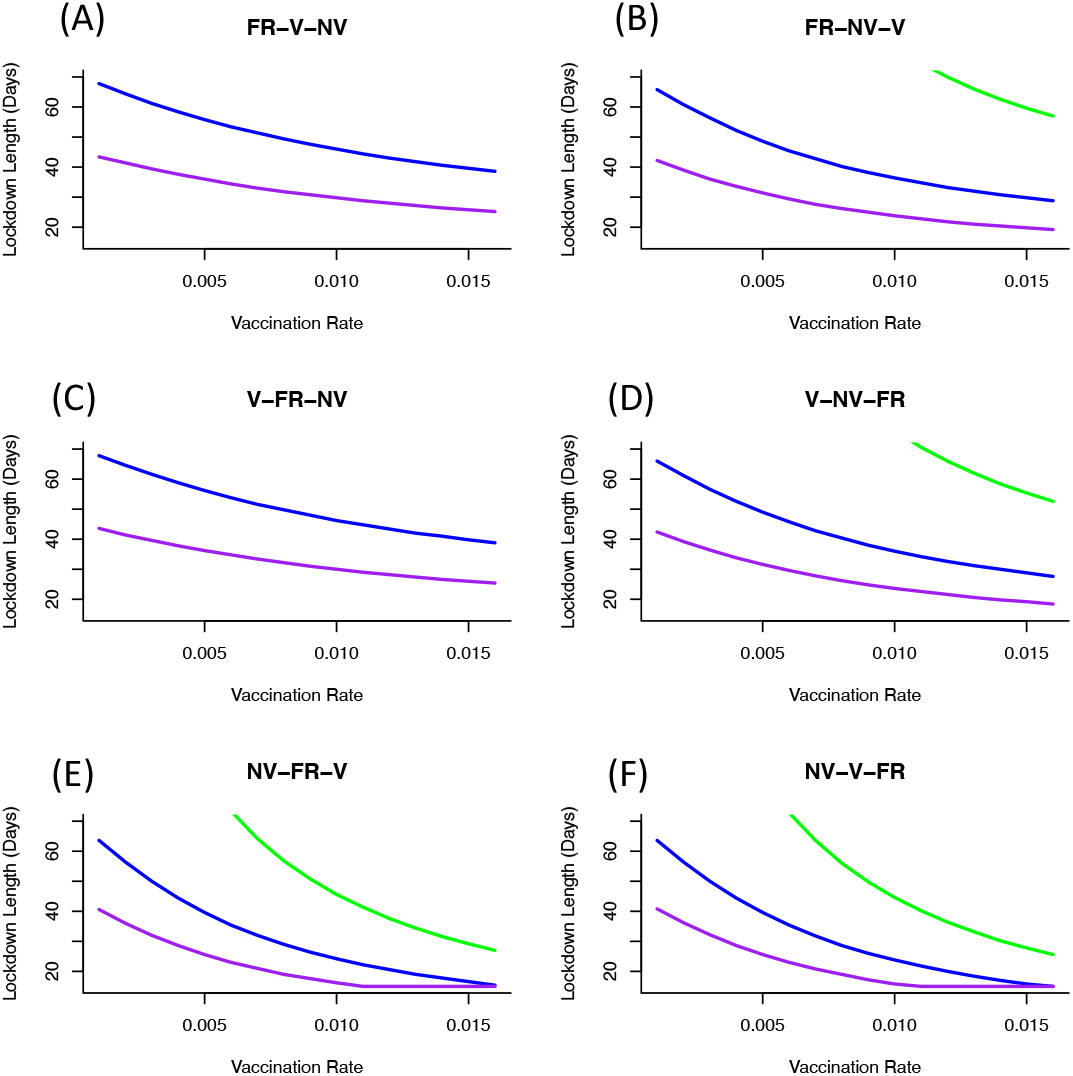
Vaccine sequence to minimize cumulative mortality (below 80K) for varying duration and severity of lockdown and vaccination rate. Sequence is to deliver vaccine to first group for 30 days, first and second group from 31-60 days and all groups after 60 days. Only moderately severe (reducing transmission by 60%) or severe (reducing transmission by 80%) achieve optimal control of mortality. Including non-vulnerable group (NV) in first or second phase of vaccination achieves better control and can reduce the severity and duration of lockdowns. If front-line workers (FR) are first in line for vaccination then the optimal sequence is shown in figure B. If vulnerable group (V) is first in line then to achieve optimal control of mortality requires vaccination of non-vulnerable group before key worker group (figure D).[(purple line - 80% reduction in *β*, blue line - 60% reduction in *β*, green line - 40% reduction in *β*]

### Vaccination demand: declining vaccine uptake rates

Vaccine demand and uptake can influence the outcome of disease mitigation and control measures. One concern is that as a substantial number of people are vaccinated, and potentially in tandem with initial declines in infection rates, then there will be an emerging complacency and vaccine adoption will fall. However, exponentially declining vaccine uptake is most likely to disrupt control measures and lead to resurgences in disease spread and increases in mortalities (Figure 6). Again, the use of lockdowns of different durations and/or severities can mitigate against this sort of loss of control. Even for weakly restrictive lockdowns (where disease transmission is only reduced by 20%) and vaccine uptake declines exponentially (here set at 2.0*x*10^−4^ per day), long duration lockdowns can reduce cumulative mortality levels (Figure 6). In fact, more severe lockdowns for this sort of loss of vaccine uptake can limit the duration of these lockdowns and lead to effective disease control.

**Figure 6:**
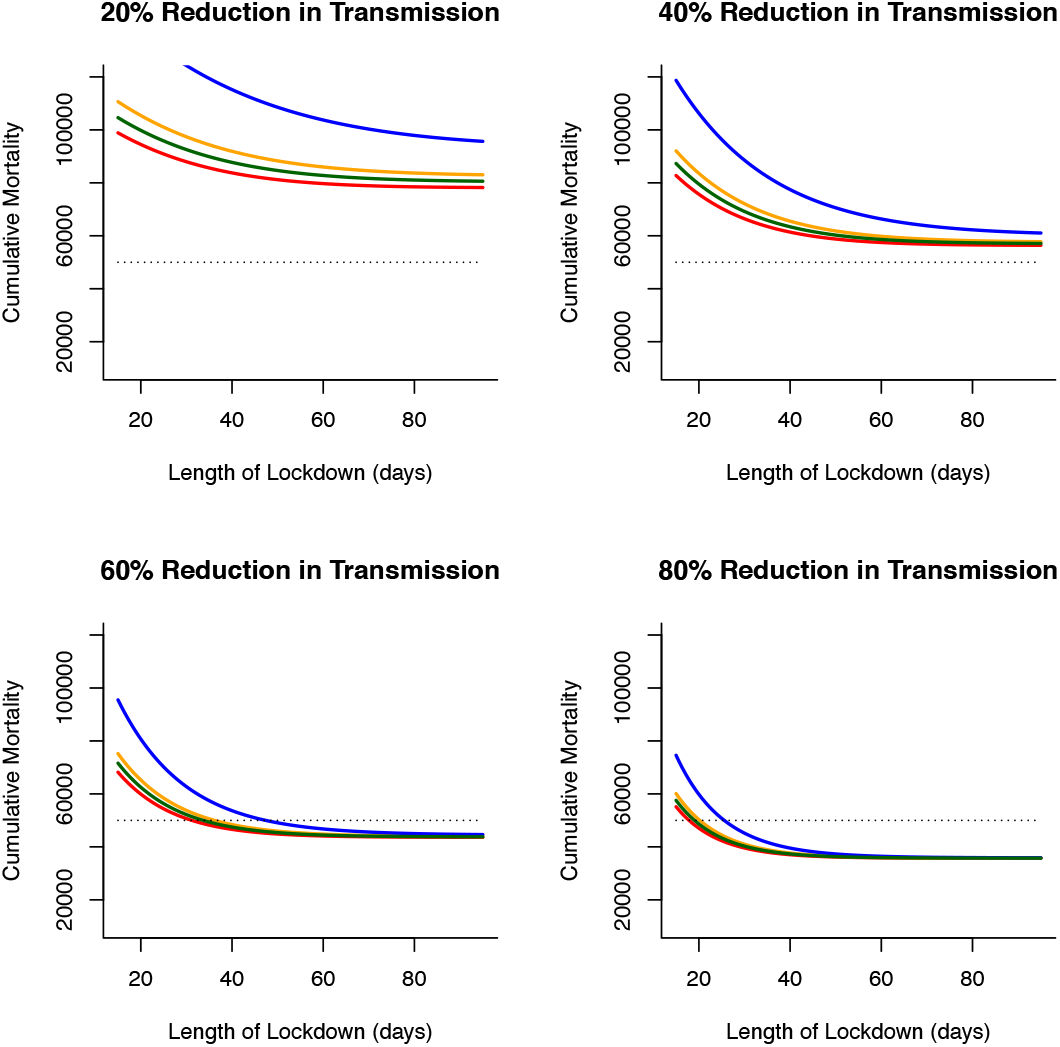
Effects of declining vaccine demand uptake and length of lockdowns (in days) on cumulative virus-induced mortality (dashed line shows 50K threshold) for different lockdown severities (as reductions in virus transmission during lockdown). Fixed (inelastic) vaccine demand always leads to the lowest levels of cumulative mortality (red line). For declining (elastic) vaccine demand, rapidly (exponential) declining vaccine uptake (blue line) leads to the largest difference from fixed vaccine (compared to linear ((orange line) or polynomial declines (green line)). The effects of elastic vaccine demand on cumulative mortality can be offset by increasing length (days) and severity (reduction in virus transmission) of lockdowns.

## Discussion

Here, using mathematical modelling approaches we have investigated how combining vaccinations and lockdowns can be used to control virus levels (in terms of disease induced levels of mortality). We show that for different levels of vaccination (or vaccine efficacy), lockdowns of different duration and/or severity can be implemented to mitigate levels of mortality. In particular, we highlight particular combinations of vaccination and lockdowns that can achieve effective virus control. Out of a number of potential exit strategies that include promoting natural herd immunity, virus elimination or releasing lockdowns as treatments become available, Sheikh et al. (2020) [17] advocate approaches whereby lockdowns are relaxed and virus infection levels are managed through tracing strategies. While this might be an achievable endgame, to reach this point requires, as we have demonstrated here, requires combinations of approaches. Rather than relaxing lockdowns as a strategy, our analysis suggests that combining them with low coverage vaccination (or low vaccine efficacy) allows optimal solutions to be found to mitigating levels of disease.

A non-random distribution of vaccinations may be ineffective even in behaviourally homogeneous populations [18]. Moreover, for heterogeneous populations, evaluating the full contact matrix to derive an appropriate vaccination strategy may be impossible. Here, as we use the simplifying approximation that within and between contacts are similar (*β*_*ji*_ = *β*), the non-random distribution of vaccines can be inefficient at achieving sufficient coverage for the disease to fade out. Similar findings for age-structured models [19] and vaccine sharing strategies (Huntingford et al. in review) corroborate this finding. Here, this occurs due to population size differences between the groups and the relative differences that this has on mitigating levels of disease-induced mortality. In particular, we find that vaccinating individuals in the vulnerable group (a relatively small group) must be rolled out together with mass vaccination across the wider larger non-vulnerable group to achieve necessary public health benefits of reducing mortality and herd immunity.

These sorts of well-planned vaccination strategies may to lead to spatial and temporal clusterings. These groupings and others such as social clusterings generate further levels of heterogeneity that achieving optimal outcomes based on vaccination strategies alone may be challenging. Groups that eschew vaccinations may make disease control more difficult. In fact, ‘clustering of exemptions’ necessitates greater vigilance around the emergence of a ‘critical mass’ where individual decisions to decline vaccination impinge on the collective (public health) benefit and restricts vaccine coverage [20, 21, 22]. Here, we account for these factors, and consider declining vaccination uptake should groups decide to alter behaviours during mass-roll out of the vaccine. As expected, rapid (exponential) decline of vaccine uptake is most precipitous in terms of optimal disease control outcomes. Our results confirm that lockdowns and other non-pharmaceutical interventions can be used to mitigate against the effects of social clusterings and loss of vaccine uptake/efficiacies. However, rapidly understanding the way in which these groupings form will be critical to determine how robust in terms of severity and duration lockdowns need to be to achieve optimal disease control outcomes for COVID-19.

Optimal control approaches have been widely used in understanding the control of infectious diseases and there have been several applications to understanding the SARS-CoV-2 pandemic. For instance, our own work [23, 24](Huntingford et al. in review) has focused on the use of optimal control approaches to understand how the use of NPIs and lockdowns could be eased so as to minimize hospitalizations, and how circuit-breakers could be optimally used to disrupt epidemic peaks, and the optimal approaches to sharing vaccines between nations. Other studies have focussed on the use of optimal control approaches for mitigating disease mortalities and the use of NPIs [25] and how vaccinations could be administered to minimize mortalities [26].

Perkins & Espana (2020) [25] use COVID-19 epidemic data from the USA to parameterise an unstructured SEIR framework with additional asymptomatic, hospitalisation and vaccinated classes to investigate the optimal use of NPIs. In their formulation, only susceptible individuals received the vaccine and optimal solutions then focused on minimizing both use of NPIs (as a reduction in disease transmission) and deaths. Perkins & Espana (2020) find that relaxing NPIs too soon can have major implications for longer term disease control and that maintaining stricter levels of control, minimizes deaths. Here, our results support this finding that the severity of control (i.e. as expressed as reductions in disease transmission *β*) can mitigate levels of disease-induced deaths. Our main finding is that in addition, with vaccination, optimal strategies to minimize deaths can, under certain conditions, offset the need for severe or long lasting NPIs.

Libotte et al. (2020) [26] use the SIR and optimal control approach to investigate vaccine delivery strategies. In their study, the objective functional is focused to minimize the number of infected individuals and the total amount of vaccine required. Their use of an inequality constraint is used to model limitations related to vaccine availability and production. With the choice of linear terms in the objective functional, the optimal solutions are on-off (bang-bang) control with variable time between delivery of vaccines to minimize the number of infections. Our results contrast with this bang-bang control. This highlights the effects of different uses of cost structures. Here, we use quadratic increasing costs to capture difficulties in achieving vaccination target as the number of vaccinated individuals increases, as well as the application of the optimal control problem to a structured (rather than unstructured) population. In our key result, the sequence of vaccine delivery to different groups is critical to achieving disease control and minimizing the public health burden of disease-induced deaths.

As noted, neither of these previous studies consider population structure and the interaction between vaccines and NPIs as concomitant approaches to disease control and minimizing disease induced deaths. We argue that these sort of optimal control approaches provide a ‘weight of evidence’ for more pluralistic approaches to controlling the infection, and especially as appropriate constraints can be included in solving numerical optimal models of the epidemiological dynamics.

In conclusion, although vaccinations for COVID-19 are starting to be approved for use, and countries are thereby implementing mass inoculation programs, many places are experiencing further waves of infection. This requires a difficult balance between providing the benefits of vaccination programs as a route to returning to normality, yet still requiring the continued use of NPIs (such as strict lockdown rules) in the interim. For this reason, it is timely to investigate the options available to minimise the time in lockdown.

Here, we have used a mathematical model to investigate the combined effect of lockdowns and during a mass vaccination plan. For the three quantities of different rates of vaccinations, extent of restrictions that impacts virus transmission and number of deaths that will protect health services, we derive the shortest lockdown length. Our mathematics of optimisation determines the shortest lockdown time for the three parameters, and critically, across different options of which groups of people to vaccinate first. We use three discrete cohorts of people, of vulnerable, front-line workers and non-vulnerable. As an additional component to our numerical calculations, we also allow for “vaccine elasticity”, where individuals may become less concerned about receiving a vaccine as disease infection rates fall. We also allow consideration of vaccines that are not fully efficacious as we evaluate optimal outcomes in terms of shortest lockdowns for prescribed maximum levels of mortality. We argue that our use of appropriately developed structured epidemiological models provide a robust way to investigate these epidemiological outcomes. Our optimal control approaches allow the best combinations (here for parameter constraints applicable to the UK) to be determined. However, these outcomes are parameter-dependent, and might change across different locations and/or temporal scales.

Our headline findings are as follows. As might be expected, to require relatively short lockdowns, NPIs have to be sufficiently severe as to suppress transmission. Less effective vaccines imply longer lockdowns, as does a larger vaccine elasticity. However, to achieve appropriate levels of disease control and contrary to the standard approach of vaccinating the most vulnerable first, we find that the optimal vaccination policy is to inoculate the larger (non-vulnerable) demographic group first, then followed by front-line workers and then the vulnerable. While this finding might appear counter-intuitive, given the order-of-magnitude difference in death rate for those encouraged to shield against COVID-19 (i.e. in the vulnerable category). The reason for this finding is our analysis assumes that the time required to vaccinate the vulnerable group is identical to that of the much larger non-vulnerable group. As the non-vulnerable group is much larger, many more people are vaccinated per day under that assumption, causing the disease to decline more quickly and yet still constraining the overall number of deaths.

We hope our analysis will encourage additional theoretical and empirically validated studies to understand further how the demographic heterogeneity (by group or age structure) impacts the epidemiological dynamics and outcomes of virus spread. Such analyses can include the associated immediate effects of lowered transmission by lockdowns, both within and between different groups, as well as the impact of choice of vaccination order on required lockdown lengths and sterilising/non-sterilising effects of vaccination in preventing onward disease transmission. In particular, the impact of choice of vaccination order on required lockdown lengths is worthy of substantial on-going investigation.

## Data Availability

The code used for all numerical analyses and simulations is available at https://osf.io/xvunt/

## Appendix

### Optimal Control

The aim is to find an way to control infection (by minimizing disease-induced mortality) through an epidemic under different transmission rates and increasing difficulty of vaccination delivery as the number of vaccinated individuals increases.

### Governing equations

We begin by introducing the governing equations (see main text for further details) We use an S-I-R-V framework to describe the epidemiological dynamics. The dynamics for susceptible individuals follow

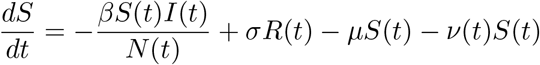

The dynamics for infected individuals follow:

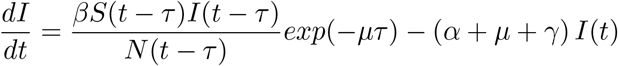

The dynamics for recovered individuals follow:

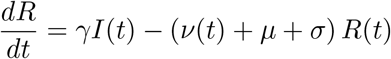

and the dynamics for the total number of vaccinated individuals are:

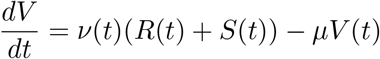

where *β* is the disease transmission rate, *σ* the loss of immunity, *µ* is the background death rate, *ν*(*t*)is the vaccination rate (on both susceptible and recovered individuals), *τ* is the incubation window, *α* is the disease induced death rate and *γ* is the disease recovery rate. N(t)=S(t)+I(t)+R(t)+V(t).

The objective functional is defined in terms of the rate of vaccination, the number of individuals vaccinated and level of disease induced mortality such that during the epidemic of time length *T* the ‘costs’ of vaccination increase. The goal is to minimize these ‘costs’ of vaccination and keep daily disease-induced mortality below a threshold (*Z*) where the control is vaccine rate (*ν*(*t*)):

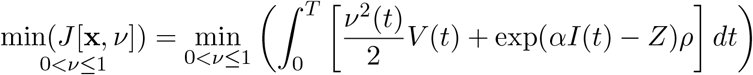

where *T* is the length of the disease epidemic wave and *Z* is the critical level of daily disease induced mortality that can not be exceeded.

To solve the optimization problem, a Hamiltonian operator (*H*) and adjoint system (see below) are formed. As with all optimal control problems, the Hamiltonian operator is formed as *H* = *f* (*t*, **x**, *ν*) + *λg*(*t*, **x**, *ν*), where *f* represents the ‘cost’ function, *g* represents the governing equations (through time *t* for dynamical system **x** and control *ν*), and *λ* is a multipler function (Kamien & Schwartz 2012). For the unstructured disease model, the Hamiltonian operator is:

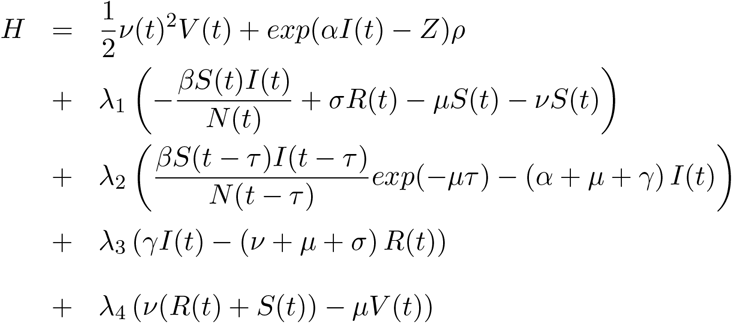

Expressions for the characterization of the control are derived from minimizing the Hamiltonian operator. Each adjoint variable (*λ*_*i*_) satisfies an equation found by differentiating the Hamiltonian operator with respect to the corresponding state variable, and then negating this derivative:

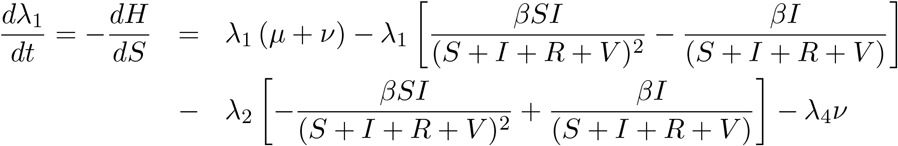

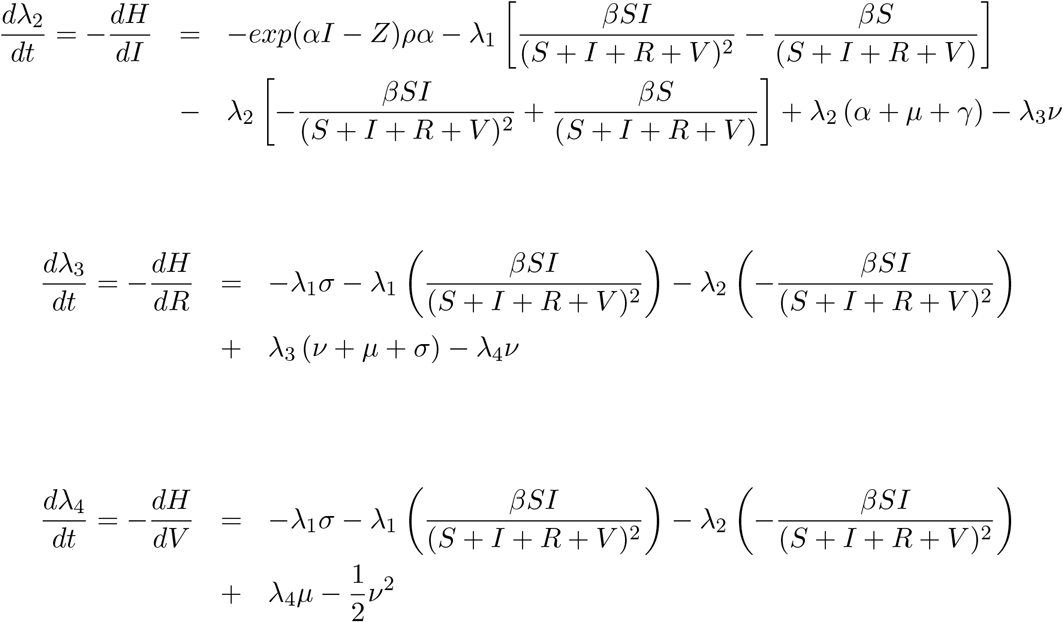

With appropriate boundary conditions, the optimal control (*ν*) then minimizes the Hamiltonian operator such that 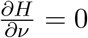 for *ν* = *ν*^*\**^:

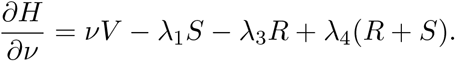

where *ν*^*\**^ is :

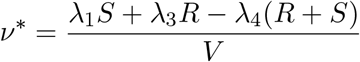

The second derivative of *H* indicates the solution is a minimum as:

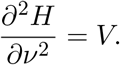

This is positive when *V* > 0, so solutions are determined as the number of vaccinated individuals increases.

